# Voxelwise Lesion Symptom Mapping in Pediatric TBI: An ENIGMA Mega-Analysis

**DOI:** 10.64898/2026.10.05.26364816

**Authors:** Emily L Dennis, Lauren Blake, Evelyn M Deutscher, Morgan Hafen, Ella M Hartsell, Elizabeth S Hovenden, Jamie Johnson, Finian Keleher, Courtney McCabe, Jake Mitchell, Emma N Read, Madi Reading, Dayna B Thayn, Robert F Asarnow, Talin Babikian, Brenda Bartnik-Olson, Erin D Bigler, Karen Caeyenberghs, Dana DeMaster, Linda Ewing-Cobbs, Christopher C Giza, Kristen R Hoskinson, Marsh Königs, Jeffrey E Max, Tricia Merkley, Mary R Newsome, Alexander Olsen, Nicholas P Ryan, Gershon Spitz, Stacy J Suskauer, Anne L Wheeler, Brandon A Zielinski, Hannah M Lindsey, Elisabeth A Wilde

**Author notes:** Correspondence to: Dr. Emily L Dennis, TBI and Concussion Center, Dept of Neurology University of Utah School of Medicine.

## Abstract

Focal lesions are common after complicated mild-to-severe traumatic brain injury (cmsTBI) in children, yet it is unclear whether the location of focal damage relates to cognitive outcome beyond overall lesion burden. Combining 13 international cohorts through the ENIGMA Pediatric cmsTBI working group, we conducted voxelwise lesion-symptom mapping (VLSM) to test whether focal lesion location relates to cognitive outcome, and how this compares with overall lesion volume. From 357 patients with imaging that passed quality assurance, 67 had a focal lesion visible on T1-weighted MRI along with at least one performance-based cognitive outcome (D-KEFS Trail Making conditions and WISC/WAIS Coding). We tested whether lesion location related to cognitive outcome using VLSM, adjusting for age, sex, and time since injury, and separately tested total lesion volume as a predictor of each outcome. Lesions in the right middle frontal gyrus were associated with poorer performance on number sequencing (D-KEFS Trail Making condition 2), a timed visuomotor sequencing task (false discovery rate corrected). In separate regression analyses, greater lesion volume was associated with poorer number-letter switching and motor speed. The switching association remained significant after adjusting for injury severity, whereas the motor-speed association did not. In the largest lesion sample assembled in pediatric cmsTBI to date, focal lesion location and injury burden index separable contributions to cognitive outcome, with number sequencing linked to focal prefrontal damage and switching and motor speed to overall lesion volume, underscoring the value of precise lesion characterization for large-scale multi-site neuroimaging.

## Introduction

Traumatic brain injury (TBI) is among the most common causes of acquired neurological disability in childhood, with an estimated annual global incidence across all severities of 226.4 per 100,000 in children and adolescents.^1^ While up to 90% of pediatric TBI cases are associated with negative clinical imaging,^2^ complicated mild-to-severe TBI (cmsTBI) is distinguished by the presence of visible intracranial pathology and carries a substantially higher risk of persisting long-term cognitive impairments. Among children with positive imaging, outcomes vary widely, with some showing lasting deficits in cognitive and functional domains while others recover fully.^3–5^ Identifying the injury features that account for this variability remains a central challenge, with direct implications for prognosis and for targeting rehabilitation.^6,7^

The lesions underlying cmsTBI are heterogeneous, reflecting the varied mechanical forces of injury. Direct impact can produce focal contusions, most often in the frontal and temporal poles, which can evolve into encephalomalacia and focal atrophy in the chronic phase.^8,9^ Rotational and shearing forces produce traumatic axonal injury, while hematomas and increased intracranial pressure can cause secondary damage.^10^ A single patient may present with a mixture of these pathologies at once. Of these, focal parenchymal lesions are the most consistently detectable on T1-weighted (T1w) MRI, whereas much of the remaining pathology is better seen on other sequences.^11^

A substantial literature has examined how focal parenchymal lesions relate to outcome, focusing largely on lesion burden. For example, greater total lesion volume, along with injury severity and lesion depth, has been associated with poorer global and cognitive outcome in both adult and pediatric cohorts.^12–15^ However, a large portion of outcome variance remains unexplained by lesion volume alone, raising the question of whether the anatomical location of focal damage carries additional, function-specific information.^16,17^

Voxelwise lesion-symptom mapping (VLSM) enables us to track the link between lesion location and outcome directly by testing at each voxel whether the presence of a lesion relates to performance on a given outcome.^18^ Since the first VLSM study,^18^ the approach has been applied most extensively in stroke, where well-defined vascular territories produce lesions that overlap reliably across patients and map onto multiple outcome domains.^18–21^ Key methodological considerations include sufficient spatial overlap of lesions to ensure statistical power, correction for multiple comparisons across voxels, and whether to include total lesion volume as a covariate. Including total lesion volume can reduce confounding from overall severity but may also remove variance that is genuinely location-specific.^22–24^

Applying VLSM in TBI is more challenging, because TBI lesions are more heterogeneous and less spatially consistent than those of stroke.^25–28^ Most prior VLSM work in TBI stems from the Vietnam Head Injury Study and comprises penetrating injury in adults, in which lesion location, defined on computed tomography, has been linked to social cognition, executive function, and neuropsychiatric symptoms.^29–34^ Penetrating TBI resembles stroke in producing relatively focal, overlapping lesions, but it is far less common than non-penetrating TBI.^35^ Evidence in adult closed-head TBI is sparser: the few voxelwise studies have localized poor outcome to lesions in deep structures such as the corpus callosum, subcortical gray matter, and brainstem, while analyses of overall injury burden have found total lesion volume to be a significant predictor of outcome.^36,37^ Pediatric studies show a similar pattern, but have characterized lesion location only at the level of coarse parcellations or lobes rather than voxelwise, and have generally found lesion volume to be more strongly associated with cognitive function than regional location.^12–15^ To our knowledge, no study has applied VLSM in a pediatric TBI cohort, likely because the required sample sizes exceed those typically available in this population.^38^

The Enhancing Neuroimaging Genetics through Meta-Analysis (ENIGMA) consortium addresses this limitation through harmonized analysis of pooled legacy data.^39^ Combining 13 cohorts within the ENIGMA Pediatric cmsTBI working group, we assembled the largest pediatric cmsTBI neuroimaging sample to date to examine how both lesion location and lesion volume relate to cognitive outcome. We analyzed lesion location using VLSM and lesion volume in separate regression analyses, treating the two as complementary analyses. We hypothesized that focal lesions would negatively correlate with cognitive performance, with the strongest associations in frontal regions given their vulnerability in TBI.

## Methods

Original studies were approved by the individual IRBs for each respective institution. All participants provided written or verbal informed assent, while parents provided written informed consent.

### Participants

Data were drawn from 13 international cohorts participating in the ENIGMA Pediatric cmsTBI Working Group. The combined sample included 498 T1-weighted (T1w) images from 357 children and adolescents with complicated mild-to-severe traumatic brain injury. During image quality control, 32 images were excluded for poor quality (motion, artifacts) and 20 images had registration errors that could not be manually recovered, leaving 446 images. Of these, 171 had no focal lesions, yielding 275 lesion-positive images. We then applied inclusion criteria for the voxelwise analysis. Of the lesion-positive images, 40 were from patients with no cognitive data shared, 26 lacked a recorded time since injury, and 130 with cognitive data but lacking the two most common tests (see below), leaving 79 images from 67 patients. Because 12 patients contributed images at more than one timepoint, and including multiple images from the same patient violates the independence assumption of the voxelwise model, we restricted the analysis to one image per patient, yielding a final sample of 67 patients (**Figure 1**). For the primary analysis we retain the earliest available scan per patient, when focal lesions are most reliably visible on T1w before chronic resorption and atrophy. We repeat the full analysis retaining the latest available scan as a sensitivity analysis to confirm that findings did not depend on timepoint selection. Sample sizes vary across outcome measures because of missing data on individual tests (range: n = 47– 67 per measure; see **Table 1**). Cohort details, including inclusion and exclusion criteria and recruitment strategy, are shown in **Supplementary Table 1**.

**Table 1.** Sample characteristics (N = 67 patients). Values are mean (SD) for continuous variables and n (%) for categorical variables. Statistics are computed on the analytic sample of 67 patients, one image per patient (earliest scan). n reflects the number of patients with available data for each measure. GCS = Glasgow Coma Scale; WISC = Wechsler Intelligence Scale for Children; WAIS = Wechsler Adult Intelligence Scale; D-KEFS = Delis-Kaplan Executive Function System.

| Characteristic | Mean (SD) | Range | n |
| --- | --- | --- | --- |
| **Demographics** |  |  |  |
| Age at scan, years | 14.14 (3.06) | 7.75–19.50 | 67 |
| Age at injury, years | 12.74 (3.98) | 0.66–18.80 | 67 |
| Sex, female | 21 (31.3%) | — | 67 |
| Time since injury, weeks | 75.11 (123.32) | 4.14–759.86 | 67 |
| GCS | 9.41 (4.01) | 3–15 | 56 |
| **Lesion characteristics** |  |  |  |
| Lesion volume, cc | 6.37 (13.40) | 0.01–85.97 | 67 |
| **Processing speed** |  |  |  |
| WISC/WAIS Coding (scaled score) | 8.61 (3.33) | 1–16 | 67 |
| **Processing speed and executive function (D-KEFS Trail Making, scaled scores)** |  |  |  |
| Condition 1 (visual scanning) | 9.96 (2.44) | 5–14 | 48 |
| Condition 2 (number sequencing) | 9.52 (3.36) | 1–15 | 54 |
| Condition 3 (letter sequencing) | 8.83 (3.92) | 1–15 | 48 |
| Condition 4 (number-letter switching) | 8.33 (3.50) | 1–15 | 48 |
| Condition 5 (motor speed) | 9.64 (2.70) | 3–14 | 53 |

**Figure 1.**
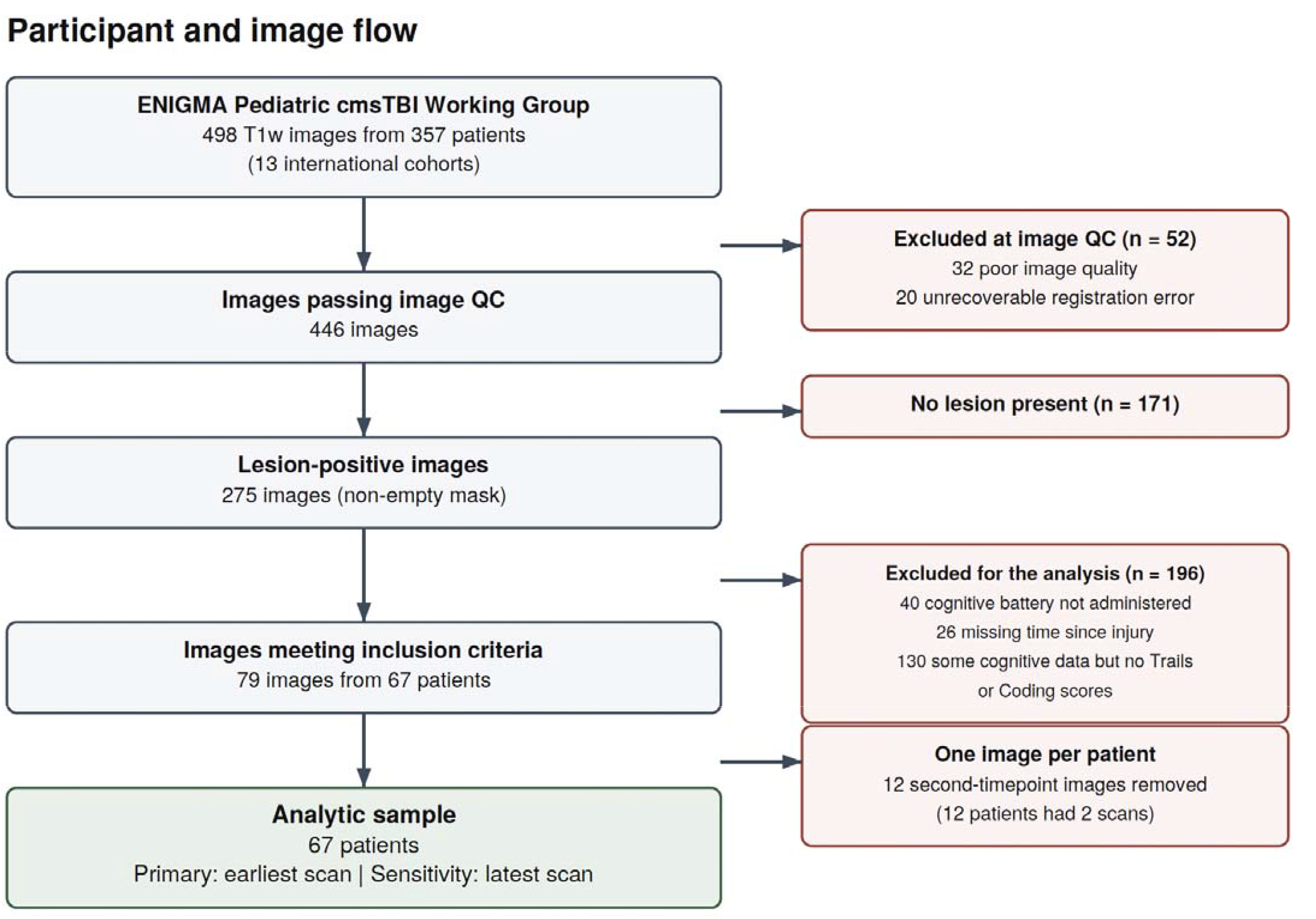
Participant analysis flow. Voxelwise analyses include six performance-based cognitive outcomes (D-KEFS Trail Making, WISC/WAIS Coding). Per-outcome samples range from 48 to 67 patients depending on test availability. For the 12 patients with two timepoints, the primary analysis retains the earliest scan; the latest scan is used in a sensitivity analysis. *Note. Voxelwise analyses include six performance-based cognitive outcomes (D-KEFS Trail Making, WISC/WAIS Coding). Per-outcome samples range from 47 to 67patients depending on test availability. For the 12 patients with two timepoints, the primary analysis retains the earliest scan: the latest scan is used in a sensitivity analysis.*

### Neuropsychological Measures

Six cognitive outcome measures were examined, all chosen as they were the most common measures collected across cohorts. Executive function and processing speed were assessed using the D-KEFS Trail Making Test (conditions 1–5) and the WISC-IV/WAIS-IV Coding subtest. The D-KEFS Trail Making Test comprises five conditions. Condition 1 (visual scanning) requires rapid cancellation of a target among distractors. Conditions 2 and 3 measure sequencing speed, connecting numbers (Condition 2) or letters (Condition 3) in order. Condition 4 (number-letter switching) requires alternating between number and letter sequences and is the primary index of cognitive flexibility, or set-shifting. Condition 5 (motor speed) measures graphomotor speed alone. Conditions 1, 2, 3, and 5 are predominantly measures of visuomotor processing speed, whereas Condition 4 additionally taxes set-shifting; comparing Condition 4 against the other conditions isolates the switching component from processing speed.^40^ In all cases, scaled scores were used, standardized for age.

### MRI Acquisition

T1-w MR images were collected at each site and shared with the University of Utah. Details on imaging parameters for each site are shown in **Supplementary Table 2**.

### Lesion Mask Preparation

Lesions were defined as focal parenchymal tissue loss, including encephalomalacia and surgical or drainage tracts. Diffuse volume loss, ventricular enlargement, and generalized atrophy were not traced, as these reflect distributed rather than focal pathology. Non-lesion abnormalities such as craniotomy defects and surgical hardware were also excluded. Binary lesion masks were manually traced on each participant’s native-space T1-w image by trained raters at the University of Utah. Dr. Evelyn Deutscher was trained by a board-certified neuroradiologist and she was responsible for training all of the 11 raters (MH, EH, ESH, JJ, FK, HML, CM, JM, ENR, MR, DBT). Training data from an independent training dataset were reviewed by two senior raters (ELD and EMD) and staff were approved to proceed to study data once they achieved a Dice similarity coefficient (DSC) of at least 0.6 against reference masks. Each lesion was segmented and reviewed in a three-stage process. First, a primary rater performed the initial manual delineation. Second, a second primary rater independently reviewed and, where necessary, edited the segmentation. Third, a senior rater (ELD, EMD, or HML) reviewed and approved the final segmentation, making minor edits where needed; any such edits were communicated to the primary raters to refine subsequent segmentations. Primary raters performed initial delineation and cross-review, while senior raters provided final review and approval before a mask entered analysis.

### Image Processing

Brain extraction was performed on the native-space T1-w images using Advanced Normalization Tools (ANTs) brain extraction (v2.5.3)^41^ with age-appropriate templates: the Nathan Kline Institute (NKI) 10 and under template for participants aged 16 years and younger,^42^ and the standard NKI template for older participants (>16 years). This approach reduces age-related masking biases, including the under-masking of pediatric brains when adult priors are applied and the over-masking of adult brains with pediatric priors are used. All brain extractions were visually inspected using a custom web-based quality control interface and failed extractions were manually corrected. Brain-extracted T1-w images were then registered to the Montreal Neurological Institute’s 152 brain 1mm template using ANTs symmetric normalization (SyN). To minimize the influence of lesioned tissue on image registration, the inverse of the binarized lesion mask was supplied to ANTs as a cost-function mask.^43^ This approach excluded lesion voxels from the similarity metric, thereby reducing lesion-related registration bias. The resulting warp fields were applied to both the T1-w and the lesion mask (using nearest-neighbor interpolation for the binary mask) to bring all data into common template space. Registration quality was visually inspected for all subjects using the same web-based system used for checking brain extractions.

### Voxelwise Lesion-Symptom Mapping

Voxelwise lesion-symptom mapping (VLSM) was conducted using Permutation Analysis of Linear Models (PALM, alpha version 119^44^) implemented in MATLAB. Analyses were restricted to voxels where at least three patients had lesion overlap, balancing the reduced power and unstable estimates at voxels with minimal overlap against the loss of spatial coverage that higher thresholds impose in a heterogeneous, moderately sized sample.^45,46^ At each voxel within this mask, a general linear model was fit with the cognitive outcome measure as the regressor of interest and covariates for age at scan, sex, and time since injury. Two contrasts tested each direction of the lesion-behavior association: contrast 1 tested for a positive relationship between outcome score and lesion presence, and contrast 2 tested for a negative relationship, the hypothesized direction in which lesion presence relates to worse performance.

Voxelwise t-statistics were assessed for significance using PALM with 10,000 permutations, threshold-free cluster enhancement (TFCE^47^), and tail approximation to improve the precision of low p-values.^48^ Family-wise error (FWE) correction was applied at the voxel level as the primary threshold. Because permutation-based FWE control can be conservative when lesion overlap is low and true effects are sparse, we additionally report false discovery rate (FDR; Benjamini-Hochberg) correction as a more sensitive complementary threshold. We considered results surviving FWE correction (p ≤ .05) as primary evidence and results surviving FDR correction (q ≤ .05) as supporting evidence, and we report both.

The primary analysis did not include total lesion volume as a covariate. Although simulation work indicates that VLSM results can be spatially misplaced and that including lesion size as a correction factor can reduce this misplacement,^46^ volume adjustment carries a competing risk in this setting. Because total lesion volume is, by definition, the sum across voxels of the same lesion masks tested in the voxelwise model, volume and voxelwise lesion status are intrinsically collinear, and adjusting for volume therefore removes variance that is itself partly location-specific rather than purely burden-related. This can suppress genuine location effects, so we treated the unadjusted analysis as primary and repeated it with total lesion volume as a covariate in a sensitivity analysis to confirm that any location associations reflected lesion location specifically rather than overall lesion burden.^22^ Total lesion volume was additionally examined as a predictor of each outcome in separate regression analyses.

Clusters were characterized by their extent in voxels, peak voxel coordinates in MNI152 1mm space, and Harvard-Oxford cortical and subcortical atlas labels at the peak and at local maxima within each cluster.

### Lesion Volume

To complement the voxelwise analysis, we tested whether total lesion volume predicted each cognitive outcome using linear regression. Lesion volume was computed for each participant by summing the number of lesioned voxels in template space from the warped binary lesion mask and converting to cubic centimeters (cc). Because the distribution of lesion volumes was positively skewed, lesion volume was log-transformed prior to analysis. For each of the six outcome measures, two linear regression models were fit. Model 1 included log-transformed lesion volume, age at scan, sex, and time since injury as predictors. Model 2 added Glasgow Coma Scale (GCS) score to assess whether any volume–outcome associations were independent of initial injury severity. Subjects with missing GCS data were excluded from Model 2 only. Statistical significance was evaluated at ⍰ = .05, uncorrected for the number of outcomes.

## Results

The analytic sample comprised 67 patients with cmsTBI, each contributing one lesion-positive image (**Table 1**). Patients were on average 14.14 years old at scan (SD 3.06, range 7.75 to 19.50) and 12.74 years at injury (SD 3.98), and 21 (31.3%) are female. Time since injury varied widely, from 4 to 760 weeks (mean 75.11, SD 123.32), reflecting the retrospective, multi-cohort design. GCS scores were available for 56 patients (mean 9.41, SD 4.01). Total lesion volume also spanned a broad range, from 0.01 to 85.97 cc (mean 6.37, SD 13.40), with the distribution skewed toward smaller lesions. Cognitive performance was near normative expectations on the simplest measures but showed reliable deficits on more demanding ones. Mean scaled scores ranged from 8.33 to 9.96 (normative mean 10, SD 3) and sample sizes ranged from 48 to 67 depending on availability of the cognitive outcome. One-sample t-tests against the normative mean were non-significant for number sequencing (Trails 2), visual scanning (Trails 1), and motor speed (Trails 5), but significant for letter sequencing (Trails 3; t(47) = −2.06, p = .045), number-letter switching (Trails 4; t(47) = −3.30, p = .002), and WISC/WAIS Coding (t(66) = −3.42, p = .001), which place greater demands on set-shifting and processing speed. The proportion of children scoring more than one standard deviation below the normative mean (scaled score < 7) exceeded the normative expectation of roughly 16% on the switching, letter sequencing, and Coding measures (25.0%, 27.1%, and 23.9%, respectively), indicating that group means near the normative average obscured a substantial subset with clinically meaningful impairment. Participants without focal lesions (from the original full set of 357 children) did not differ on any cognitive outcome from participants with focal lesions (all p > .20; WISC/WAIS Coding raw and scaled, D-KEFS Trails 1-5).

### Lesion Overlap

A voxelwise lesion overlap map was computed by summing all binarized, template-space lesion masks (**Figure 2**). Maximum overlap was 14 subjects (21%). Lesions were most frequent at the temporal poles, with the greatest overlap on the ventral and anterior temporal surface (9%), and at the frontal poles, concentrated ventromedially in orbitofrontal and paracingulate cortex and extending to the midline superior frontal cortex (21%). Callosal involvement centered on the genu and anterior body (7%).

**Figure 2.**
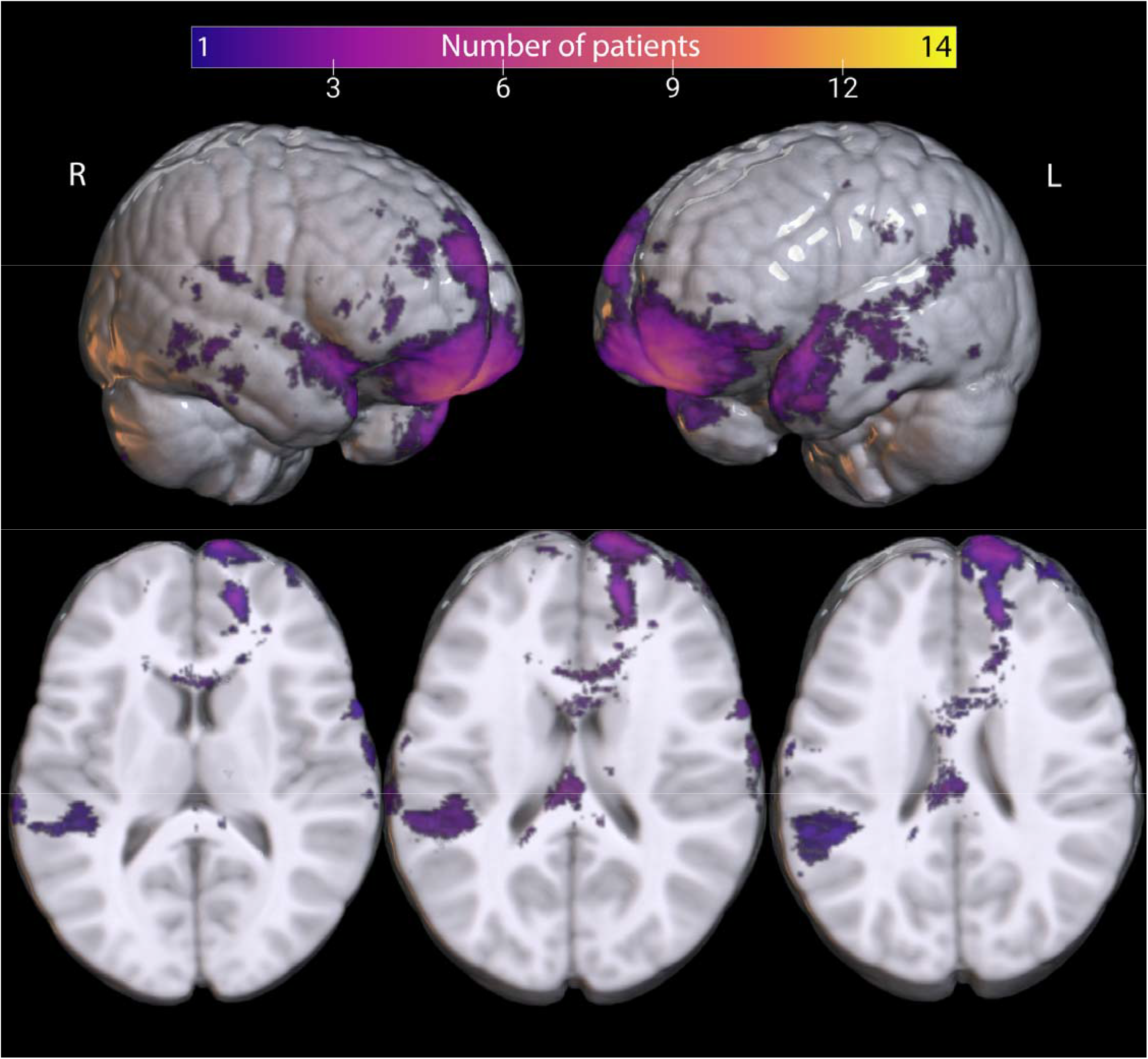
Lesion overlap. Binarized lesion maps were collapsed to show where lesion distribution overlapped between patients, with number of patients corresponding to the color bar. The maximum overlap was 14.

### Voxelwise Lesion-Symptom Mapping

We report FWE correction as the primary, more stringent threshold, controlling the probability of any false positive across voxels, alongside FDR correction as a more sensitive secondary threshold, controlling the expected proportion of false positives among significant voxels. No voxel or cluster survives FWE correction for any of the six cognitive outcomes. One association survives false discovery rate correction: for D-KEFS Trail Making condition 2 (number sequencing), lesion presence relates to worse performance in the earliest-scan analysis (*q* = .040), with the peak in right middle frontal gyrus (MNI 30, 36, 32, **Table 2, Figure 3**). The peak lies in the cortex rather than in the region of highest lesion overlap. The number-sequencing cluster, while surviving FDR correction, overlapped the lesions of only 4 patients, reflecting the low spatial overlap achievable even in a sample of this size. This finding should therefore be interpreted as a spatially specific but low-prevalence association, and replication in larger samples is needed.

**Table 2.**
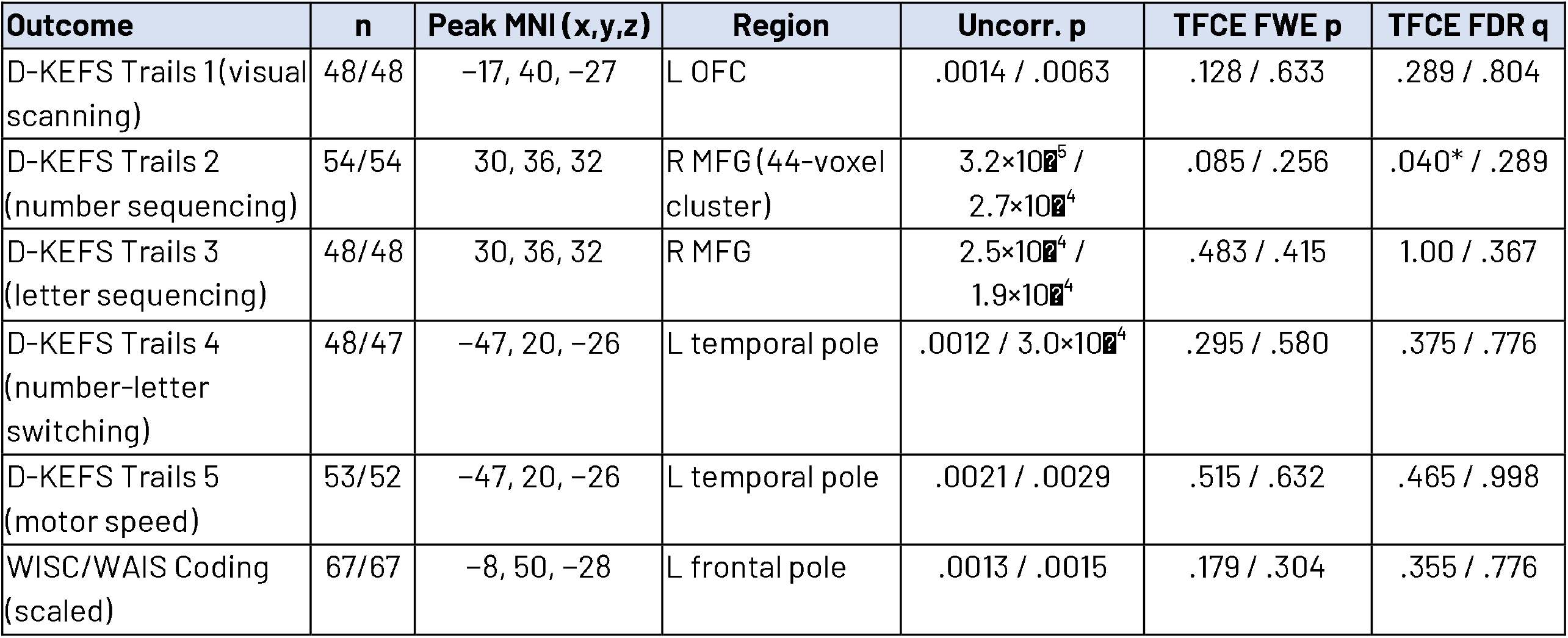
VLSM results. Voxelwise lesion-symptom mapping across six neurobehavioral outcomes for the hypothesized contrast (lesion presence associated with worse performance) in the primary analysis (no lesion-volume covariate). Analyses used PALM with 10,000 permutations, threshold-free cluster enhancement, and tail acceleration. No outcome survives family-wise error correction. All models include age at scan, sex, and time since injury. OFC = orbitofrontal cortex, MFG = middle frontal gyrus.

| Outcome | n | Peak MNI (x,y,z) | Region | Uncorr. p | TFCE FWE p | TFCE FDR q |
| --- | --- | --- | --- | --- | --- | --- |
| D-KEFS Trails 1 (visual scanning) | 48/48 | -17, 40, -27 | L OFC | .0014 / .0063 | .128 / .633 | .289 / .804 |
| D-KEFS Trails 2 (number sequencing) | 54/54 | 30, 36, 32 | R MFG (44-voxel cluster) | $3.2 \times 10^{-5}$ / $2.7 \times 10^{-4}$ | .085 / .256 | .040* / .289 |
| D-KEFS Trails 3 (letter sequencing) | 48/48 | 30, 36, 32 | R MFG | $2.5 \times 10^{-4}$ / $1.9 \times 10^{-4}$ | .483 / .415 | 1.00 / .367 |
| D-KEFS Trails 4 (number-letter switching) | 48/47 | -47, 20, -26 | L temporal pole | .0012 / $3.0 \times 10^{-4}$ | .295 / .580 | .375 / .776 |
| D-KEFS Trails 5 (motor speed) | 53/52 | -47, 20, -26 | L temporal pole | .0021 / .0029 | .515 / .632 | .465 / .998 |
| WISC/WAIS Coding (scaled) | 67/67 | -8, 50, -28 | L frontal pole | .0013 / .0015 | .179 / .304 | .355 / .776 |

**Figure 3.**
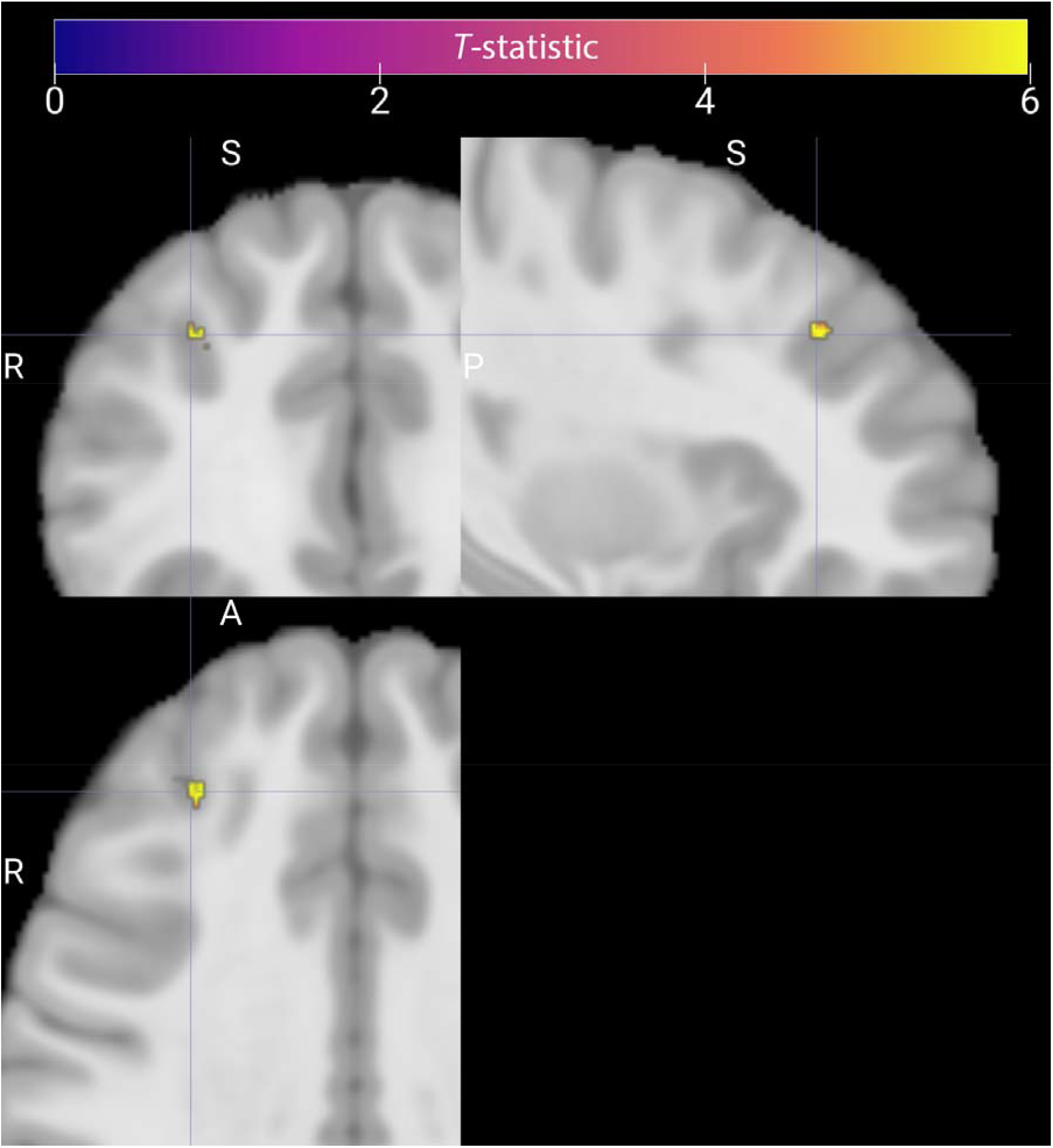
Voxelwise lesion-symptom mapping result for D-KEFS Trail Making number sequencing (Condition 2). Lesion presence is associated with worse performance in a single cluster in right middle frontal gyrus extending into frontal pole (peak MNI 30, 36, 32; 44 voxels), shown in coronal, sagittal, and axial views through the peak, overlaid on the NIHPD pediatric template. Color reflects the voxelwise t-statistic within the cluster surviving false discovery rate correction (q = .040). No cluster survives family-wise error correction. Crosshairs mark the peak voxel; images follow radiological convention (R = right).

**Figure 4.**
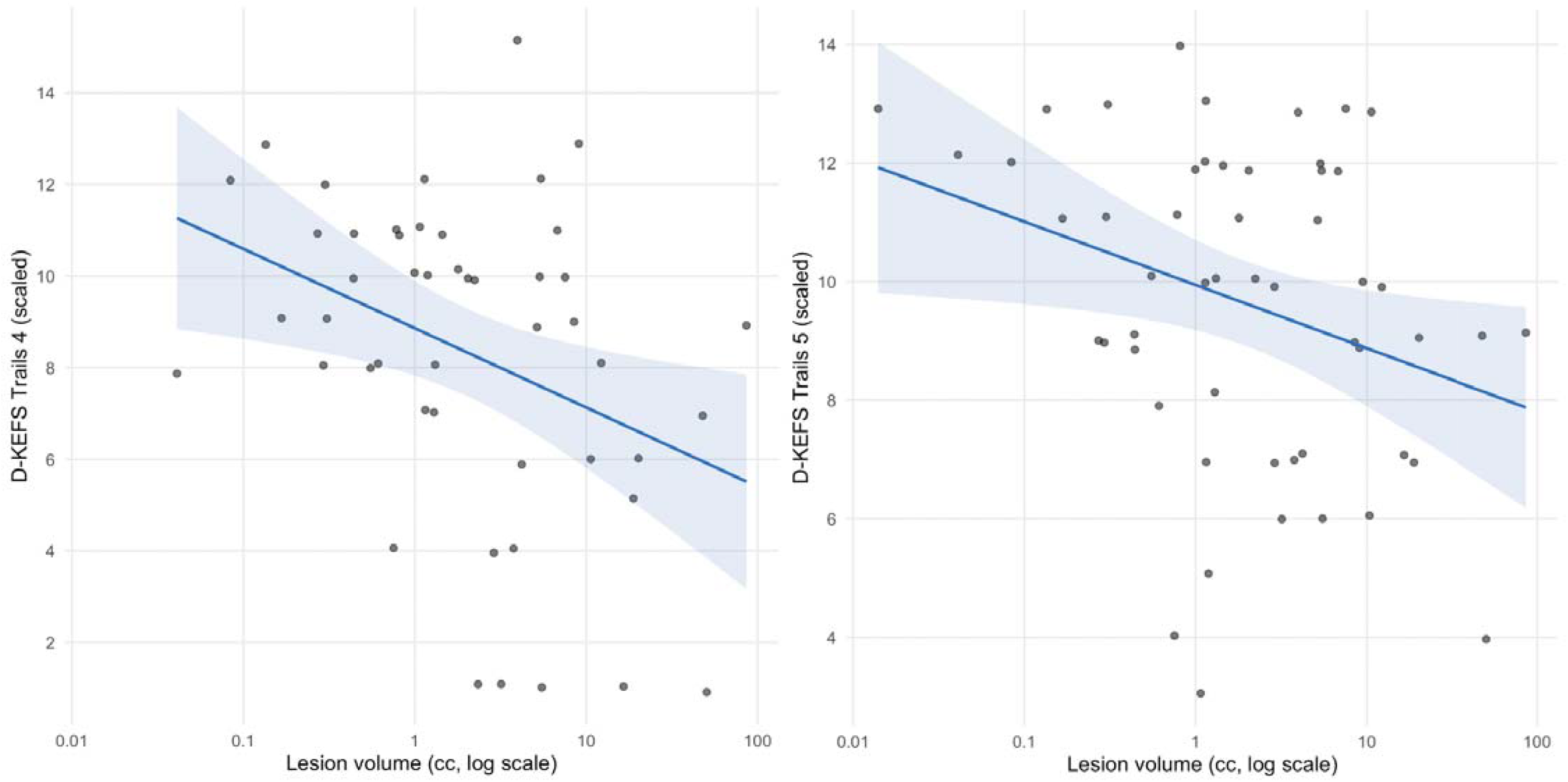
Associations between lesion volume and cognitive outcomes. Total lesion volume in relation to D-KEFS Trail Making number-letter switching (Condition 4, left) and motor speed (Condition 5, right), shown as age-scaled scores (normative mean 10, SD 3). Lesion volume is plotted on a log scale. Points are individual participants; the line shows the linear fit with 95% confidence band. Associations are unadjusted, and only the Condition 4 association remained significant after adjustment for injury severity (Glasgow Coma Scale score).

Across the remaining five outcomes, no association survives correction in either timepoint analysis. At uncorrected thresholds, peak associations are in the hypothesized direction (lesion presence relating to worse performance) for all six outcomes, with peaks distributed across right middle frontal gyrus, left orbitofrontal and frontal polar cortex, and left temporal pole (**Table 2**). Peaks in the opposite direction converge on a single right frontal periventricular white-matter location (near MNI 18, 51, 6) affected in a large proportion of the sample, consistent with lesion-density bias rather than functional localization.

Additional sensitivity analyses may be found in **Supplementary Note 1**.

### Total lesion volume and cognitive outcomes

In regression models adjusting for age, sex, and time since injury, larger total lesion volume relates to worse performance on two executive measures at the earliest timepoint: D-KEFS number-letter switching (⍰ = −0.70, *p* = .033, Table 3) and motor speed (⍰ = −0.49, *p* = .032). Both associations are in the expected direction, with larger lesions predicting lower scaled scores. Adjusting additionally for GCS score, in the subset with this measure available, leaves the switching association essentially unchanged (⍰ = −0.79, *p* = .037) and attenuates the motor-speed association (*p* = .18). Neither association survives correction for the six outcomes tested, and both associations attenuate at the latest timepoint (*p* = .069 and .096). No other outcome relates to lesion volume in either model or at either timepoint.

**Table 3.** Lesion volume results. Associations between total lesion volume and cognitive outcomes (N = 67 patients). Ordinary least-squares regression of each cognitive outcome on log-transformed total lesion volume, in the primary sample (earliest scan per patient). ⍰ is the standardized coefficient for log lesion volume; negative values indicate that larger lesions are associated with worse performance. Model 1 adjusts for age at scan, sex, and time since injury; Model 2 additionally adjusts for Glasgow Coma Scale score, in the subset with GCS available. *p < .05, uncorrected. No association survives correction for the six outcomes tested. At the latest timepoint, the Trails 4 and Trails 5 associations attenuate to trends (p = .069 and .096, respectively, Model 1).

|  | Model 1: primary (age, sex, TSI) |  |  | Model 2: + GCS |  |  |
| --- | --- | --- | --- | --- | --- | --- |
| Outcome | n | $\beta$ (log vol) | p | n | $\beta$ (log vol) | p |
| D-KEFS Trails 1 (visual scanning) | 48 | −0.14 | .530 | 41 | −0.05 | .845 |
| D-KEFS Trails 2 (number sequencing) | 54 | −0.14 | .623 | 43 | −0.04 | .915 |
| D-KEFS Trails 3 (letter sequencing) | 48 | −0.08 | .842 | 40 | −0.07 | .862 |
| D-KEFS Trails 4 (number-letter switching) | 48 | −0.70 | .033* | 40 | −0.79 | .037* |
| D-KEFS Trails 5 (motor speed) | 53 | −0.49 | .032* | 43 | −0.39 | .177 |
| WISC/WAIS Coding (scaled) | 67 | −0.24 | .356 | 56 | −0.27 | .340 |

## Discussion

This study investigated whether the location or overall volume of focal lesions contributes to cognitive outcomes following pediatric cmsTBI using VLSM in the largest pediatric lesion sample assembled for this purpose to date. Here we interpret three primary findings: the distribution of focal lesions across the cohort, the association between right prefrontal damage and number sequencing, and the relationship between total lesion volume and switching and motor speed. We then consider the clinical and methodological implications of applying VLSM in this population.

Focal lesions were distributed widely across the cohort but showed only modest overlap, with a maximum of 14 patients (21%) sharing a lesion at the single densest voxel. The greatest overlap fell on the ventral and anterior temporal surface at the temporal poles, with further concentration ventromedially at the frontal poles, in orbitofrontal and paracingulate cortex extending to the midline superior frontal cortex, and along the genu and anterior body of the corpus callosum. This anterior, ventral, and midline distribution is consistent with lesion patterns reported in other brain injury studies, reflecting the biomechanics of closed-head injury and the vulnerability of orbitofrontal and anterior temporal tissue against the skull base.^49^ That a sample of this size yielded a maximum overlap of only 14 patients illustrates the spatial heterogeneity of focal injury in pediatric cmsTBI, which sets an intrinsic limit on the statistical power available at any single voxel.

Our central finding is an association between focal damage to the right middle frontal gyrus (MFG), extending toward the frontal pole, and poorer performance on the D-KEFS Trail Making Test (TMT) number sequencing (Condition 2). The right MFG lies within the dorsolateral prefrontal cortex and contributes to the visual search, sustained attention, and speeded visuomotor control that timed sequencing demands. The region has also been linked to higher-order number processing: resting-state work associates numeracy with right MFG connectivity and literacy with the left MFG,^50^ and children with mathematical disabilities show altered connectivity of a frontoparietal network that includes the right MFG.^51^ We interpret our association cautiously in this respect, however, because number sequencing is the most basic TMT condition and, on factor-analytic grounds, indexes visuomotor processing speed more than the numerical competence or set-shifting that distinguish other conditions.^40,52^ Right MFG damage is therefore most likely to impair the speeded, attentionally demanding components of the task rather than number processing or higher-order executive control per se. The association emerged for a single outcome, and the sample size limits power to establish its specificity, but the peak is spatially stable, falling at the same location in the primary and supplementary analyses. We read it as evidence that focal lesion location accounts for a specific portion of cognitive outcome after pediatric cmsTBI.

A separate regression of total lesion volume, which indexes overall injury burden rather than its spatial location, implicated different outcomes. Larger lesion volume was associated with poorer performance on D-KEFS TMT number-letter switching (Condition 4) and motor speed (Condition 5). These diverged after adjustment for injury severity - the switching association remained with GCS in the model, indicating a contribution of structural burden beyond global severity, whereas the motor-speed association did not, pointing to closer dependence on overall severity. That motor speed tracked total volume rather than lesion location is consistent with its known reliance on efficient, distributed processing^53^ and with the broad vulnerability of white matter to the diffuse pathology common in pediatric cmsTBI,^54–56^ though we did not test tract involvement and do not interpret these conditions in terms of specific structural pathways. These findings align with Bourke et al. (2022), who reported that reduced brain volume, rather than the presence of focal lesions, was associated with slower processing speed in pediatric TBI.^57^

That location and volume implicated different measures is itself informative. The number-sequencing association showed no relationship with lesion volume (p = .62), and the volume associations for switching and motor speed showed no spatial localization. This double dissociation indicates that these measures relate to distinct lesion properties, and it is why we analyzed location and volume separately rather than entering volume as a covariate in the VLSM. A sensitivity analysis supports that choice: a direction-reversed association appeared only when lesion volume was covaried, the signature of a suppressor effect that arises when a covariate is strongly collinear with the spatial predictor.^46^ Covarying for volume in this setting removes genuine signal and introduces artifactual reversals rather than clarifying localization.

Neither lesion location nor lesion volume was associated with WISC/WAIS Coding performance, a measure of processing speed,^58,59^ in any analysis. Coding draws on a broad network spanning frontoparietal, motor, cerebellar, and visual regions.^60–64^ This wide recruitment, relative to the more circumscribed demands of the Trail Making conditions, may reduce its sensitivity to the effect of any single focal lesion.

### Limitations

Three limitations temper these findings. First, the analytic sample of 67 patients meets minimum standards for VLSM, but the low lesion overlap limits power. We note, however, that this sample was drawn from the largest pediatric cmsTBI lesion dataset compiled to date and meets published minimum-sample recommendations for VLSM,^65^ representing a realistic ceiling for manually segmented multi-site work in this population rather than an easily remedied shortfall. Second, the cohort spans a broad developmental range. Executive functions and their supporting white matter continue to mature through adolescence,^66,67^ and TBI can permanently alter neurodevelopment in children.^3,57,68–71^ Because of these, the functional impact of focal lesions may vary by developmental stage with rapidly developing functions potentially at greater risk for long-term impairment^72^ than well-established skills.^73,74^ Age was included as a covariate, but the sample does not support age-stratified analysis. Third, lesions were defined on T1-weighted images alone, which capture macroscopic tissue loss but not the diffuse axonal injury (DAI) that occurs in up to 60% of severe TBI.^75^ This is relevant as DAI may contribute to the burden-dependent associations we observed for switching and motor speed, which tracked total lesion volume rather than location. As diffusion, susceptibility-weighted, and FLAIR imaging capture aspects of injury that T1w does not, focal T1w lesion location represents only one facet of pathology, and combining VLSM with these modalities in future work would give a fuller account of how focal and diffuse pathology jointly shape long-term outcome.^76^

## Conclusions

Combining 13 international cohorts, we assembled the largest pediatric cmsTBI neuroimaging dataset for lesion analysis to date, and applied VLSM for the first time in this population. The scale of the assembled sample is a strength, but it also illustrates how much attrition VLSM imposes in this population. Because outcome measures varied across cohorts, we restricted analysis to the neuropsychological tests administered consistently across studies, the D-KEFS Trail Making conditions and WISC/WAIS Coding, rather than to the full battery any single cohort collected. Of 357 patients with usable imaging, 67 had a focal lesion together with at least one of these shared outcomes, exceeding the minimum recommendations for VLSM, and the covariates required for analysis. Within this analyzable sample, focal lesion location was associated with one outcome after multiple comparisons correction, performance on D-KEFS Trail Making number sequencing, localized to right middle frontal gyrus extending into the frontal pole. The remaining cognitive measures showed no lesion-location association surviving correction. Rather than a simple null result, we read this pattern as informative about the method itself. Lesion location carries clinical signal where a function has a relatively focal and consistent structural basis, but most outcomes after pediatric TBI depend on distributed networks, diffuse white matter organization, and developmental and environmental factors that focal lesion mapping is poorly positioned to capture. The single surviving association, for a processing-speed-loaded sequencing task rather than a measure of higher-order executive control, is consistent with this reading.

A practical implication follows for study design. If associations this circumscribed emerge even after assembling the largest pediatric cmsTBI imaging sample to date, and if the analyzable subset is nonetheless modest once lesion presence, outcome availability, and covariates are required, then smaller studies are unlikely to be adequately powered for VLSM unless they target a function with a strong prior expectation of focal localization. For most questions about cognition and behavior after pediatric TBI, quantitative measures of white matter microstructure and network connectivity may be more sensitive than lesion location alone, and the two are best combined.

The limited clinical yield of lesion location should not be mistaken for methodological irrelevance. Focal lesions bias intensity-based registration and confound voxelwise analyses whether or not they are the variable of interest, so they must be accounted for even when the analytic target lies elsewhere. This is the motivation for the AIMS-TBI Challenge (MICCAI), now in its third year, which supports the development of automated lesion segmentation methods.^28^ Robust, scalable lesion detection is a prerequisite for valid multi-site neuroimaging analysis in this population, independent of whether lesion location itself proves clinically predictive.

### Transparency, Rigor, and Reproducibility

This study used a retrospective, cross-sectional design drawing on existing data from 13 international cohorts. Inclusion and exclusion criteria are fully specified in the Supplement, and all exclusions are reported (**Figure 1**). Sample sizes per outcome ranged from 47 to 67 due to missing data on individual tests, reported transparently in Table 1. Sex and age at scan were included as covariates in all analyses, along with time since injury. Exploratory sex-specific analyses were not conducted given sample size constraints. Lesion masks were manually traced by trained raters using a standardized protocol implemented across all sites. Raters were approved to proceed only after achieving a Dice similarity coefficient of ≥0.6 on shared training data reviewed by expert raters. A three-stage review process was applied to all segmentations, with expert approval required before any mask entered analysis. Raters performing initial segmentation were not blinded to clinical information, which is a limitation inherent to the multi-site retrospective design; the multi-stage review process was implemented to mitigate individual rater bias. Because lesion status was determined by the presence of a non-empty warped mask on review, images without a usable lesion after registration were excluded and are reported in the participant flow (**Figure 1**). Analyses were restricted to one image per patient; for patients scanned at more than one timepoint, the earliest scan was retained for the primary analysis and the latest scan was used in a sensitivity analysis. Voxelwise analyses used permutation-based inference (PALM, 10,000 permutations) with Threshold-Free Cluster Enhancement and family-wise error correction. False discovery rate–corrected maps were generated as an additional correction, and uncorrected peaks are reported to convey the direction and strength of subthreshold effects. All contrast directions were pre-specified based on the scoring direction of each outcome measure. The primary analysis did not include lesion volume as a covariate, consistent with published recommendations for VLSM; a lesion-volume-controlled sensitivity analysis is reported, with direction-reversed findings transparently described and interpreted as suppressor effects. Total lesion volume was additionally examined in separate regression analyses. This study was not pre-registered. Data sharing follows the policies of the ENIGMA consortium and the individual data use agreements governing each contributing cohort. Group-level results and lesion overlap maps are available upon reasonable request. The lesion registration and VLSM pipeline will be made publicly available later in 2026 following completion of beta testing. FSL PALM is freely available at https://fsl.fmrib.ox.ac.uk/fsl/fslwiki/PALM.

## Supporting information

STROBE

## Data Availability

Data sharing follows the policies of the ENIGMA consortium and the individual data use agreements governing each contributing cohort. Group-level results and lesion overlap maps are available upon reasonable request.

## Acknowledgements

R01NS05400; R01NS122184; R61NS120249; K01HD083459; U54EB020403; R01MH116147; R56AG058854; P41EB015922; R01MH111671; K12HD001097; R01NS100973; R01HD061504; R01HD088438; K23HD06161; UL1TR001079; 1S10OD021648; DoD contract W81XWH-18-1-0413; NHMRC Ideas Grant 1184403; James J and Sue Femino Foundation; Hanson-Thorell Research Scholarship; UCLA Easton Clinic for Brain Health; Stan and Patty Silver; UCLA Brain Injury Research Center; KC is supported by a National Health and Medical Research Council Career Development Fellowship; DJS is supported by the SAMRC; this work was supported by The Liaison Committee between the Central Norway Regional Health Authority (RHA) and the Norwegian University of Science and Technology (NTNU) (2020/39645).

## Supplementary Material

**Supplementary Table 1.**
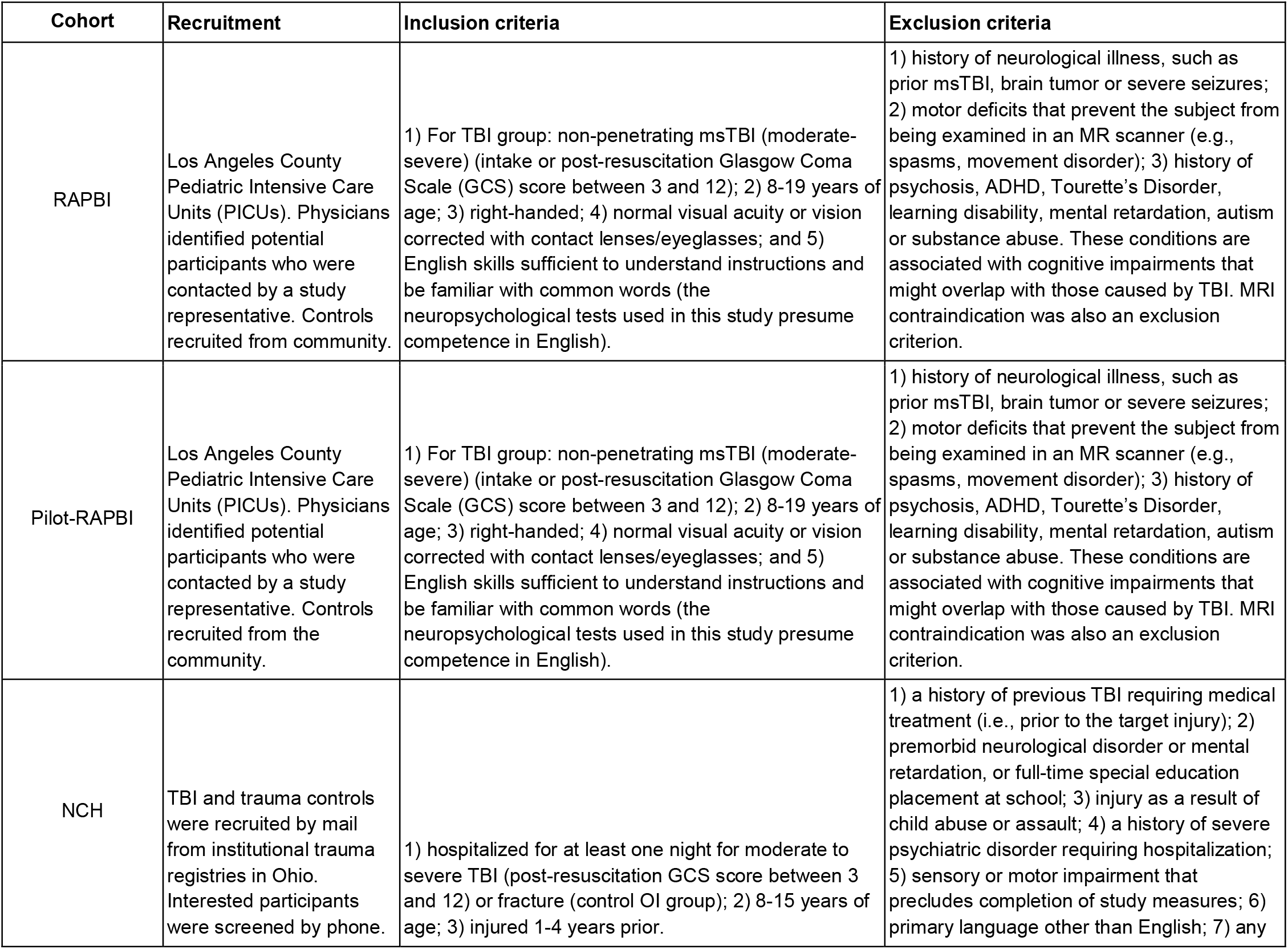
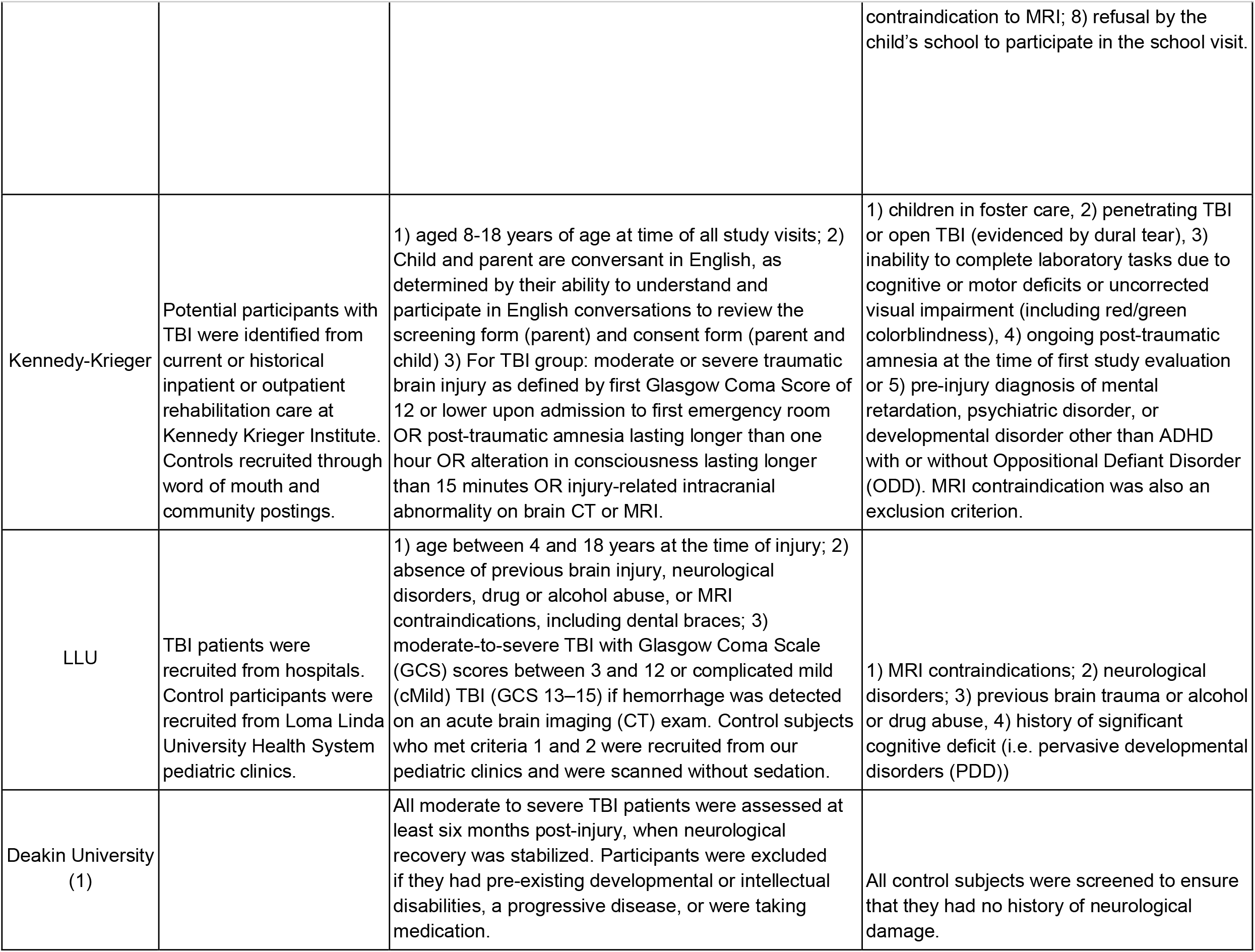
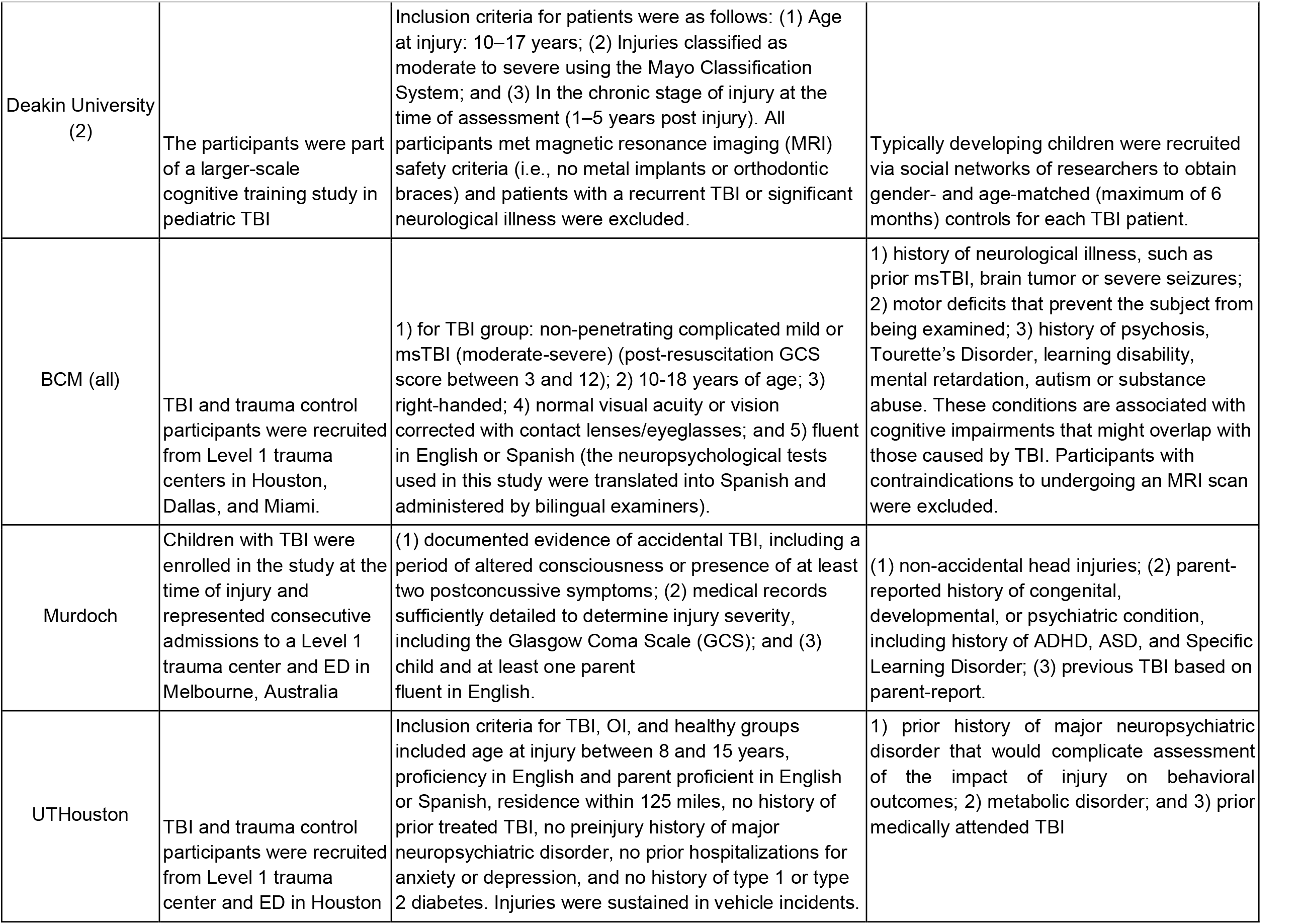
Cohort details. For each cohort, the recruitment location is shown, along with inclusion and exclusion criteria. * = cohorts with OI controls.

**Supplementary Table 2.** Scanner details. For each cohort the scanner manufacturer and model are listed, along with field strength and voxel size (in mm).

| **Cohort** | **Scanner** | **Field strength (T)** | **Voxel size (mm)** |
| --- | --- | --- | --- |
| RAPBI | Siemens TrioTim | 3T | 1x1x1 |
| Pilot-RAPBI | Siemens Sonata | 1.5T | 1x1x1 |
| NCH | Siemens Prisma | 3T | 1x1x1 |
| Kennedy Krieger | Philips | 3T | 1x1x1 |
| LLU | Siemens TrioTim | 3T | 1x1x1 |
| Deakin1 | Siemens Magnetom Trio | 3T | 1x1x1 |
| Deakin2 | Philips Intera | 3T | 1x1x1.1 |
| BCM1 | Philips Intera or Achieva | 1.5T | 1x1x1 |
| BCM2 | Philips Intera | 3T | 1x1x1 |
| BCM3 | Philips Achieva | 3T | 1x1x1 |
| Murdoch | Siemens TrioTim | 3T | 1x0.5x0.5 |
| UTHouston | Philips | 3T | 1x1x1 |

**Supplementary Figure 1.**
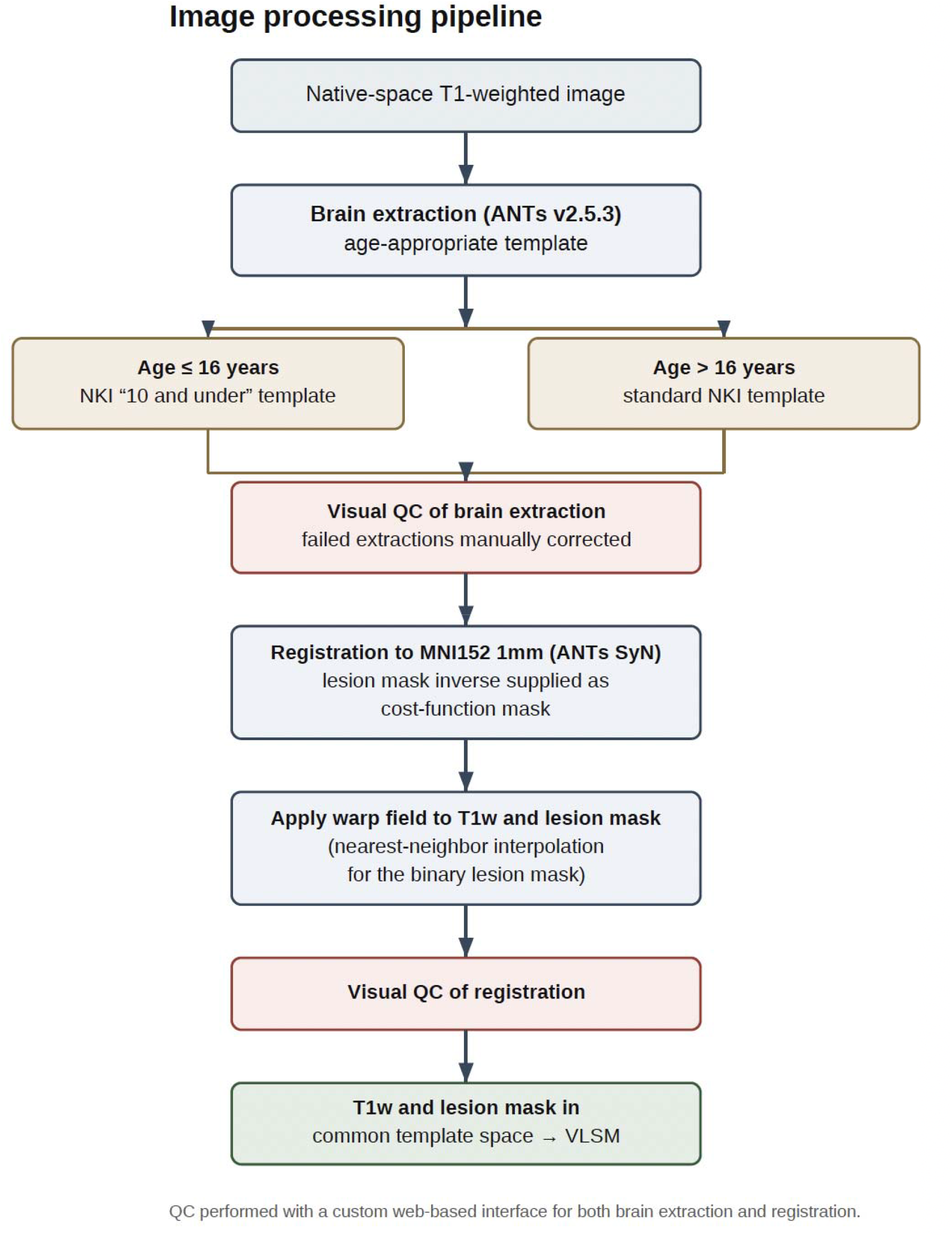
Analytical pipeline.

## Supplementary Note 1. Supplementary Results

*Voxelwise Lesion-Symptom Mapping*

While the primary analyses used the first available timepoint for the subset of participants with longitudinal data, as a sensitivity analysis we repeated analyses using the latest timepoint. Results were consistent, with the location of the peak identical in the latest-scan analysis, although the association did not survive correction at that timepoint (*q* = .289). The uncorrected peak is consistent in direction and magnitude across timepoints (earliest *p* = 3.2 × 10⍰^5^, latest *p* = 2.7 × 10⍰^4^).

*Sensitivity analysis with lesion volume covariate*

Including total lesion volume as a covariate confirms that the number-sequencing association reflects lesion location rather than overall burden, as it remains significant after adjustment (*q* = .048, earliest scan). This model produced one additional association for number-letter switching, but in the direction opposite to hypothesis and only when lesion volume was modeled (*q* = .021; nonsignificant in the primary analysis, *q* = .361).

